# Risk factors for the development of hepatocellular carcinoma (HCC) in chronic hepatitis B virus (HBV) infection: a systematic review and meta-analysis

**DOI:** 10.1101/2020.08.21.20179234

**Authors:** Cori Campbell, Tingyan Wang, Anna McNaughton, Eleanor Barnes, Philippa C Matthews

## Abstract

**Background:** Hepatocellular carcinoma (HCC) is one of the leading contributors to cancer mortality worldwide and is the largest cause of death in individuals with chronic hepatitis B virus (HBV) infection. It is not certain how the presence of other metabolic factors and comorbidities influences HCC risk in HBV. Therefore we performed a systematic review and meta-analysis to seek evidence for significant associations.

**Methods:** MEDLINE, Embase and Web of Science databases were searched from 1^st^ January 2000 to 24^th^ June 2020 for English studies investigating associations of metabolic factors and comorbidities with HCC risk in individuals with chronic HBV infection. We extracted data for meta-analysis and report pooled effect estimates from a fixed-effects model. Pooled estimates from a random-effects model were also generated if significant heterogeneity was present.

**Results:** We identified 40 observational studies reporting on associations of diabetes mellitus, hypertension, dyslipiaemia and obesity with HCC risk. Meta-analysis was possible for only diabetes mellitus due to the limited number of studies. Diabetes mellitus was associated with > 25% increase in hazards of HCC (fixed effects Hazards Ratio [HR] 1.26, 95% CI 1.20–1.32, random effects HR 1.36, 95% CI 1.23–1.49). This association was attenuated towards the null in sensitivity analysis restricted to studies adjusted for metformin use.

**Conclusions:** In adults with chronic HBV infection, diabetes mellitus is a significant risk factor for HCC, but further investigation of how antidiabetic drug use and glycaemic control influence this association is needed. Enhanced screening of individuals with HBV and diabetes may be warranted.

## Introduction

Hepatitis B virus (HBV) is a hepatotropic virus responsible for substantial morbidity and mortality worldwide. Infection can be acute or chronic, with most of the HBV disease burden attributable to chronic disease. The World Health Organisation (WHO) estimated a chronic HBV (CHB) global prevalence of 257 million for 2015, with 887,000 HBV-attributable deaths reported in the same year (1), making HBV the second highest viral cause of daily deaths (with first being the agent of the global COVID-19 pandemic, SARS-CoV-2) (2,3) of which the burden has increased in recent decades (4).

Most CHB deaths are due to primary liver cancer and cirrhosis; these conditions were responsible for over 40% of all viral hepatitis-attributable deaths. A GBD study on the global HCC burden reported a 42% increase in incident cases of HCC attributable to chronic infection between 1990 and 2015 (5), among which CHB infection was the largest contributor, responsible for more than 30% of incident cases in 2015 (5).

Multiple risk factors for HCC in CHB-infected individuals have been established, including sex, age, cirrhosis, and co-infection with human immunodeficiency virus (HIV) or other hepatitis viruses (including hepatitis C and D). Previous studies have investigated associations of comorbidities, such as diabetes mellitus (DM) (8–11) and hypertension (12,13), with risk of HCC in the general population, and The European Association for the Study of the Liver (EASL) recognises DM as a risk factor for HCC in CHB (7). As the global prevalence of comorbidities such as DM (14), renal disease(15), hypertension (16) and coronary heart disease (CHD) (17) continues to rise, these conditions are increasingly relevant to the development of HCC.

Various risk scores have been developed to predict HCC risk: the PAGE-B risk score was developed to predict HCC risk in Caucasian patients on antiviral treatment (18), while the REACH-B (19,20), GAG-HCC (21,22) and CU-HCC (21,23–25) risk scores apply to in untreated Asian populations. Existing risk scores use age, sex and HBV DNA VL to predict risk. CU-HCC and GAG-HCC include parameters for cirrhosis, however no score accounts for comorbidities such as DM or hypertension. It is possible that risk prediction could be improved by accounting for these comorbid conditions.

Despite rises in the global prevalence of relevant comorbidities, evidence concerning associations of comorbidities with HCC risk in CHB is poor and heterogeneous. Neither EASL, American Association of the Study of Liver Disease (AASLD) (33) or Asian Pacific Association for the Study of the Liver (APASL) (32) guidelines for HBV management include recommendations for enhanced screening or DM management in CHB, despite recent clinical interest in the potential utility of metformin in preventing and treating various cancers (26,27). Furthermore, there are few studies investigating associations of other potentially relevant comorbidities (such as hypertension, CHD and renal disease) and their metabolic risk factors (such as obesity and dyslipidaemia) with HCC risk. Therefore, we undertook a systematic review, aiming to summarise and critically appraise studies investigating associations of relevant comorbidities and metabolic factors with risk of HCC in CHB-infected individuals.

## Methods

### Search strategy and selection criteria

In June 2020 we systematically searched three databases (Web of Science, Embase and MEDLINE) in accordance with PRISMA guidelines (28); search terms are listed in Table S1. We searched all databases from 1^st^ January 2000 until 24^th^ June 2020, without application of any restrictions for study design applied to search terms or results, but including only full-text human studies published in English.

We combined and deduplicated search results from the three databases, prior to screening for eligibility. We excluded articles not investigating associations of comorbidities with risk of HCC and/or not restricted to CHB-infected participants. We also searched reference lists of relevant systematic reviews/meta-analyses and studies identified for inclusion to identify additional studies for inclusion. Search terms were constructed and agreed on by three authors (PM, TW and CC) and articles were screened and selected by one author (CC).

### 2.2 Data extraction and statistical analysis

One author (CC) extracted the following summary characteristics from included studies: country, publication year, study design, follow-up period, comorbidities investigated, number of participants, number of HCC cases, sex, age at baseline, risk ratio and covariates adjusted for.

We carried out meta-analysis in R (version 3.5.1) using the “meta” package (version 4.12–0) (29), including only hazard ratios (HRs) minimally adjusting for age and sex. We calculated pooled summary effect estimates using the inverse-variance weighting of HRs on the natural logarithmic scale, and quantified between-study heterogeneity using the I^2^ statistic; significance of heterogeneity was investigated using Cochran’s Q test *(p* threshold = 0.05). Where I^2^ was > 0 and heterogeneity was significant, we present both fixed- and random-effects summary estimates. We undertook multiple sensitivity analyses whereby analyses were restricted to studies adjusting for various additional confounders and for DM treatment, and stratified by DM type, in order to investigate robustness of observed associations.

### Definitions

For diabetes, we considered diagnoses of type 1 and type 2 diabetes, as well as unspecified diabetes mellitus, for pooling the effect, followed by further stratification by subtypes of diabetes if enough studies were eligible. Hypertension (HT), was defined by either a diagnosis of HT recorded as part of the medical history or current health assessment, or a measurement with mean arterial pressure (MAP) above a specified threshold. Obesity was based on BMI values, by referring the cut-off in the included studies, where 25, 27, 30 kg/m^2^ were the common threshold values used. CVD was defined broadly as an umbrella term including any of the following disease subtypes: ischeamic heart disease (IHD)/coronary heart disease(CHD), cerebrovascular disease. Dyslipidaemia was defined according to serum lipid concentrations above a certain threshold (thresholds may vary depending on healthcare setting).

### Quality appraisal

We used the Newcastle-Ottawa Scale (NOS) to assess the quality of non-randomised studies, including cohort and case-control studies (30), judging studies based on points awarded for selection of study groups, comparability of groups, and exposure/outcome ascertainment. Studies with scores of < 5, 5–7 and > 7 points were considered to be of low, sufficient and high quality, respectively.

## Results

### Study characteristics

In total our search identified 1,814 articles (899 from MEDLINE, 407 from Embase and 508 from Web of Science) (Figure 1). After deduplication, we screened 1,136 individual articles by title/abstract, from which 140 full texts were identified for full-text assessment. After exclusion of ineligible articles and reference list searching of relevant articles, we identified 40 articles for inclusion in this review. Summary characteristics of included studies are reported in Table S2.

**Figure 1.**
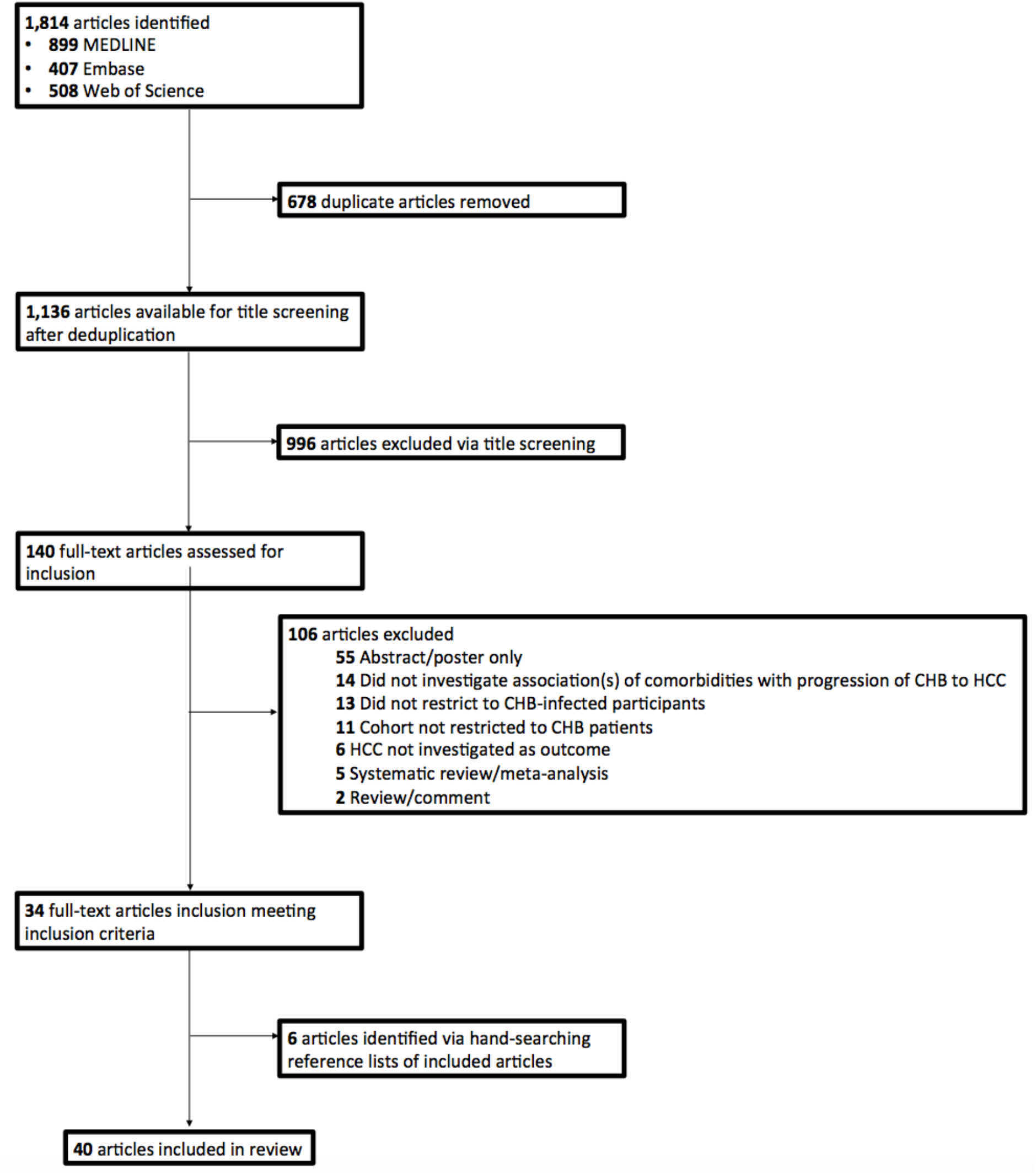
Flow chart of study selection. MEDLINE, Embase and Web of Science databases were systematically searched using relevant terms to identify relevant human studies published in English from 1^st^ January 2000 to 24^th^June 20

All studies were observational in design, with 33 cohort and 7 case-control studies included (Table S2). Thirty-two studies were conducted in Asian countries. Four studies were restricted to male cohorts and 36 were undertaken in mixed-sex cohorts. All studies recruited participants from health centres, healthcare or prescription databases, or pre-existing cohorts or cancer screening programmes. All studies were undertaken in adults, with mean/median ages of cohorts ranging between 40 and 65 years in 33 studies. Thirty-three studies investigated DM/insulin resistance/fasting serum glucose, 11 studies investigated blood hypertension/blood pressure, 7 investigated dyslipidaemia, 5 investigated obesity and cardiovascular disease. Less than 5 studies investigated other factors including renal disease, statin use and use of antidiabetic drugs.

In the 40 studies including 536,456 adults, > 30,500 HCC events occurred (we are unable to report an exact number, because one study did not report a precise number of HCC cases (31)). Sample sizes of cohort studies varied widely, ranging from 102 to 214,167 (median 3,090), with corresponding numbers of HCC cases ranged from 7 (arising among 102 participants) to 11,241 (arising among 214,167 participants). Case-control sample sizes ranged from 182 to 6,275 (median 1,122) with corresponding numbers of HCC cases ranged from 73 (out of 182 participants) and 1105 (out of 6,275 participants).

### Quality assessment

Among 40 studies, 39 had quality scores ≥5 (Tables S3 and S4). All cohort studies were of sufficient quality with 13 of these being scored as high-quality. Six case-control studies were of sufficient quality and one of poor quality. Inclusion criteria varied widely and therefore study populations were heterogeneous. In most studies, exposures and outcomes were ascertained using health assessment, imaging or record linkage. Twenty-three cohort studies and 7 case-control studies accounted for age and sex. HCC typically arise after long durations of infection, and therefore prolonged follow up allows for detection of more HCC events; among 23 cohort studies identified, only 5 cohort studies had lengths of follow-up ≥ 10 years).

### Association of diabetes mellitus with HCC risk

Thirty-six studies investigated the association of DM with risk of CHB progression to HCC, comprising 7 case-control studies (Table 1a) and 29 cohort studies (Table 1b). Four studies were restricted to males and the others included both sexes (Table S2). Mean ages at baseline in all studies were ≥40 years, respectively. Study populations were heterogeneous with variable inclusion criteria, and definitions of DM were not consistent between studies. Four case-control and four cohort studies investigated type 2 DM/insulin resistance, three case-control and seven cohort studies investigated unspecified DM, and one case-control and three cohort studies investigated both type 1 and 2 DM as a composite potential risk factor.

**Table 1a.** Effect estimates for case-control studies investigating the association of diabetes mellitus with hepatocellular carcinoma risk.

| Country, Author (Year) | Participants, n | HCC Cases, n | Age at baseline, years | Sex (% male) | Source of controls | Relative risk measure<sup>†</sup> | Risk Ratio (95% Confidence intervals) |
| --- | --- | --- | --- | --- | --- | --- | --- |
| Taiwan, Shyu (2019) (68) | 5,932 | 731 | 40-90 (range) | 56.7 | Same insurance database as cases | aHR | 1.35 (1.16–1.57) |
| US, Kennedy (2018) (36) | 1,101 | 278 | 64 | 78 | Same clinical data repository as cases | aOR | 0.6 (0.4-0.9) |
| China, Li (2018) (69) | 322 | 112 | 52 (median, HCC cases),<br>51 (median, non-HCC controls) | 18.8 (HCC cases)<br>26.2 (non-HCC controls) | Hospital controls | aOR | 2.042 (1.150-5.018) |
| China, Han (2017) (70) | 182 | 73 | 56.18 (mean) | 79.1 | Hospital controls | N/A | N/A |
| China, Gao (2013) (35) | 370 | 122 | 54.7 (mean) | 86.9 | Hospital controls | aOR‡ | 0.376 (0.175-0.807) (compensated cirrhosis)<br>0.192 (0.054-0.679) (decompensated cirrhosis) |
| China, Li (2012) (34) | 6275 | 1105 | 53.8 (mean, cases)<br>44.9 (mean, controls) | 84.7 (cases)<br>73.8 (controls) | Hospital controls | aOR | 0.8 (0.6-1.1) |
| Taiwan, Chao (2011) (37) | 1142 | 124 | 30-65 | 100 | Control group is random sample of non-HCC participants from total cohort | aHR | Those with homeostasis model assessment-insulin resistance (HOMA-IR, fasting insulin (μU/ml) X fasting glucose (mmol/l)/22.5) index scores of <0.46, 0.78-1.22 and >1.22 had HRs of 1.48 (0.90-2.44), 0.92 (0.54-1.55) and 1.96 (1.23-3.10) as compared to those with a HOMA-IR index of 0.46-0.77 |

aHR, adjusted Hazards Ratio; aOR, adjusted Odds Ratio; N/A, not available.
† Adjusted risk ratios are minimally adjusted for age and sex.
‡ Adjusted for age but not sex.

**Table 1b.**
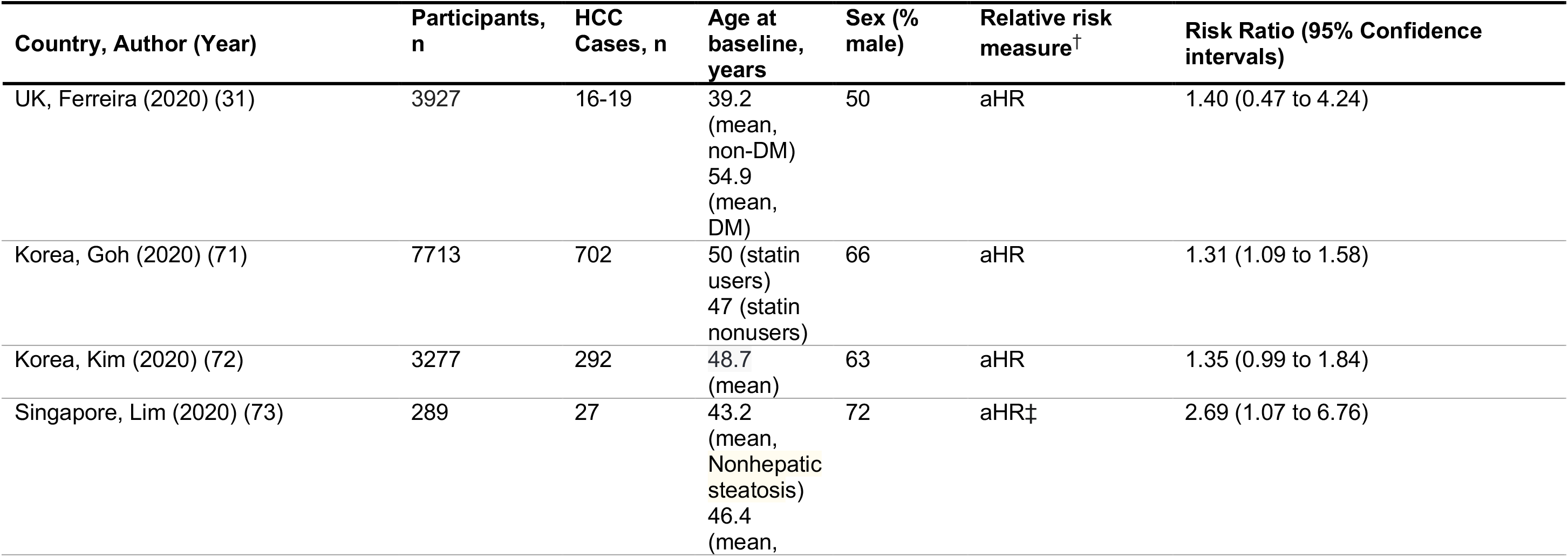
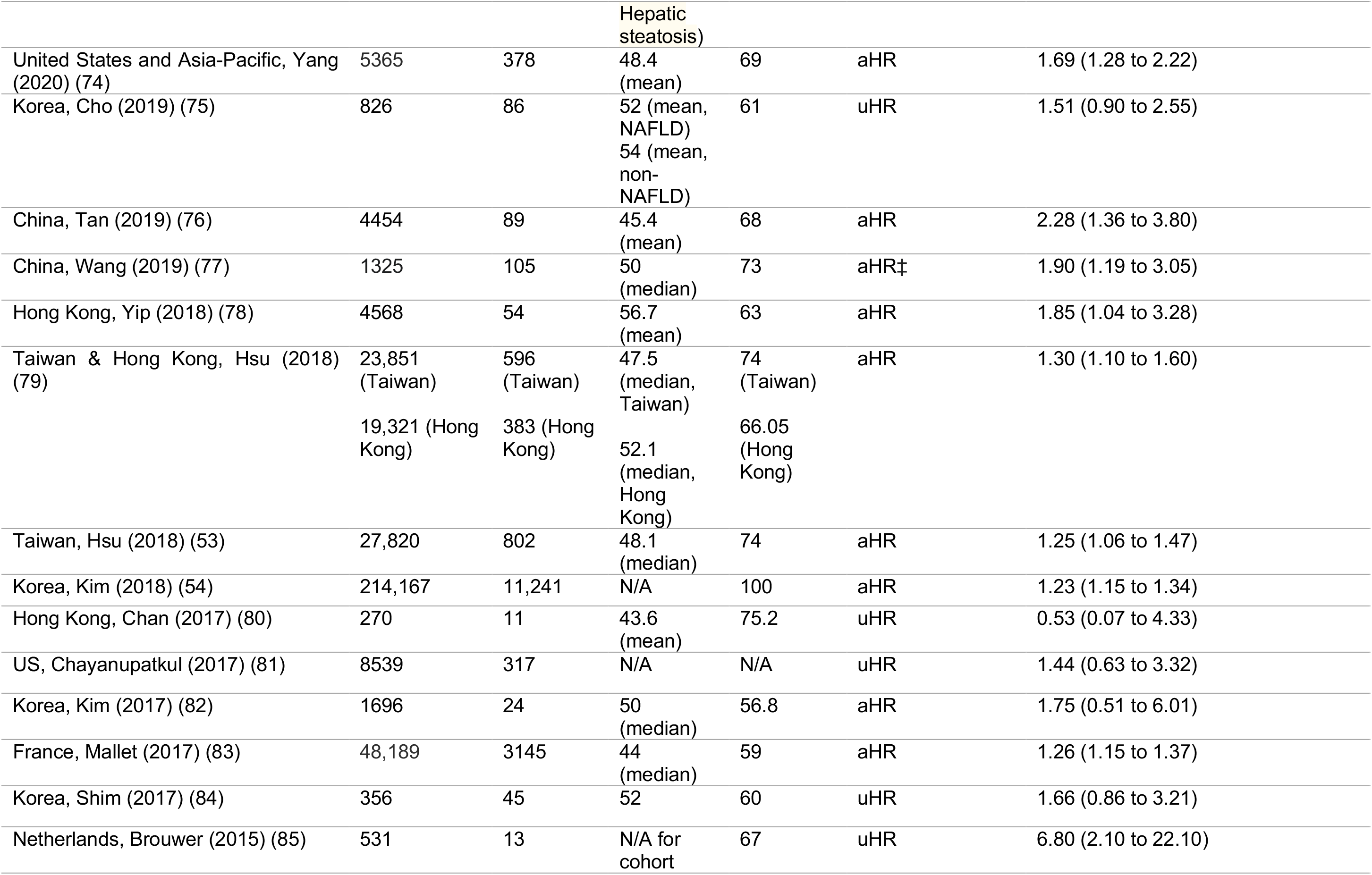
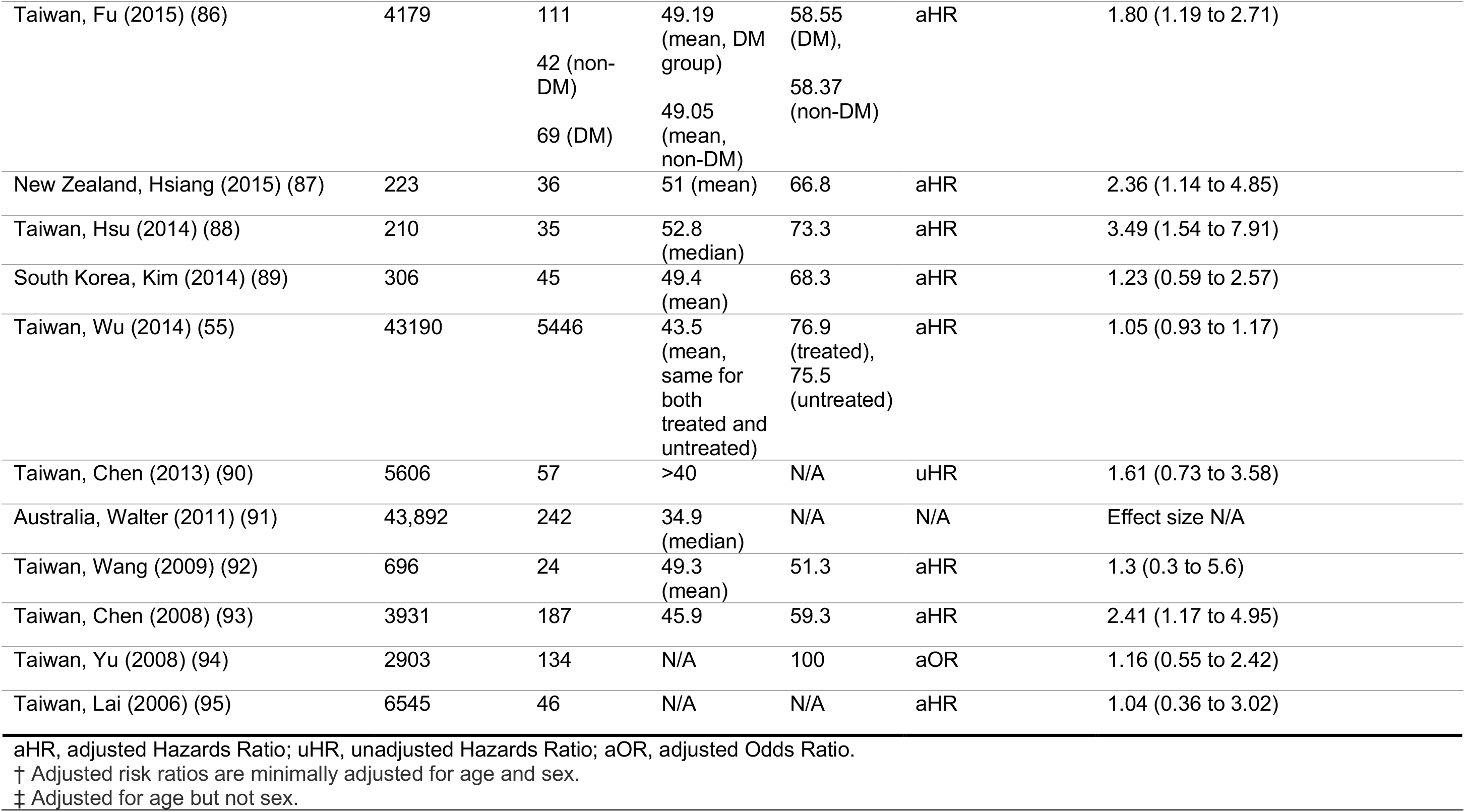
Effect estimates for cohort studies investigating the association of diabetes mellitus with hepatocellular carcinoma risk.

Of the 7 case-control studies that reported effect estimates, there was directional inconsistency between effect estimates reported in case-control studies, with 4 studies reporting an increased risk of HCC in those with DM as compared to those without, 3 studies reporting a decreased risk of HCC in those with DM, and one study failing to provide an effect estimate. Risk ratios (RRs) > 1 ranged from 1.35 to 2.04, and all were statistically significant. RRs < 1 ranged from 0.19 to 0.80, of which two were statistically significant. Among 28 cohort studies providing effect estimates (27 HRs and 1 OR), there was directional consistency with 27 of the reported RRs > 1. Effect sizes > 1 ranged from 1.05 to 6.80, with 15 RRs being statistically significant. The single RR that was < 1 was nonsignificant.

Minimal adjustment for confounders differed between case-control and cohort studies. Most case-control studies adjusted for age, sex, HCV coinfection, HIV coinfection and cirrhosis. Twenty cohort studies minimally adjusted for age and sex. Of these, 15 adjusted for HCV coinfection, 13 for cirrhosis, 12 for antiviral treatment, 10 for HIV coinfection, 9 for alcohol consumption, 7 each for HBV viral DNA load and cigarette smoking and 6 for other liver disease (including alcoholic liver disease). Eight studies excluded participants who developed HCC within the first 3 to 12 months of follow-up in their main analyses. One study did so in sensitivity analysis and found this did not modify associations observed.

### Meta-analysis of cohort studies

DM was associated with an increased risk of progression to HCC by meta-analysis restricted to HRs minimally adjusted for age and sex (Figure 2). As there was significant heterogeneity (I^2^ = 49%, *p*< 0.01), results from both fixed- and random-effects analyses are presented. In random-effects analysis risk of HCC was 36% (summary RR 1.36; 95% CI 1.23–1.49) significantly higher in DM compared to non-DM.

**Figure 2.**
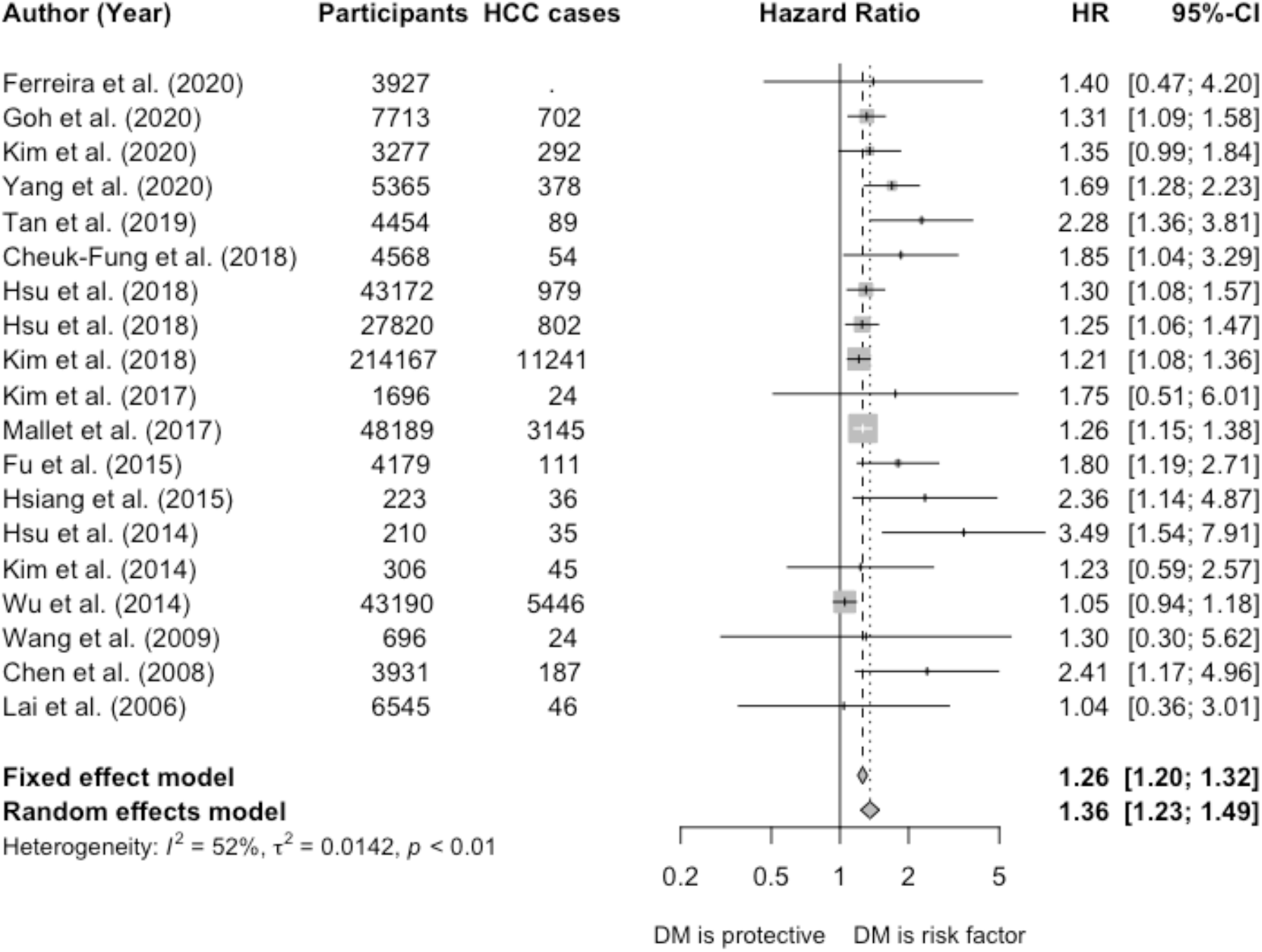
Forest plot of hazard ratios from longitudinal cohort studies investigating the association of diabetes mellitus with risk of progression of chronic hepatitis B infection to hepatocellular carcinoma. The study by Yu et al (94) provided an Odds Ratio and was excluded from the meta-analysis. Dashed vertical lines represent HR based on meta-analysis of all studies by fixed effect and random effects models. The studies for pooling the HR had sufficient quality (quality scores ≥5). HR, hazard ratio; CI, confidence interval; DM, diabetes mellitus.

### Subgroup and sensitivity analyse

We performed sensitivity analyses in order to investigate the robustness of pooled estimates to additional adjustment for HCV or HIV coinfection, cirrhosis, and DM treatment. After restricting meta-analysis to 16 studies adjusting for HCV coinfection in addition to age and sex (Figure S1), pooled HRs did not change materially. Considering 8 studies adjusting for HIV and antiviral treatment (Figure S2), pooled HR from the fixed-effects analysis was attenuated towards the null slightly but still remained significant. To investigate the robustness of the association of DM with HCC to adjustment for cirrhosis, a potential mediator, we restricted meta-analysis to studies adjusting for cirrhosis (Figure S3). This did not change pooled HRs materially.

To investigate heterogeneity between type 2 DM and unspecified DM, sensitivity analysis was performed whereby studies were stratified by DM type. Amongst studies investigating type 2 DM, heterogeneity was 33% (p = 0.18) (Figure S4). However the HR did not differ materially to that observed in the primary meta-analysis.

The association of DM with HCC risk was attenuated towards the null in studies that adjusted for metformin use, with risk of HCC 16% higher in DM participants as compared to non-DM (random effects HR 1.16, 95% CI 1.04–1.29) in analysis restricted to studies adjusting for metformin use (Figure S5). After restricting to studies adjusting to DM treatment, pooled HRs remained statistically significant.

### Association of hypertension with hepatocellular carcinoma risk

Eleven studies investigated the association of HT with risk of CHB progression to HCC, one case-control study and 10 cohort studies (Table 2). All studies were mixed-sex samples in which mean/median age at baseline was ≥40 years (Table S2). Definitions of HT were heterogeneous; most studies ascertained hypertension via record linkage, but others used health assessment or interview. Few studies defined clinical thresholds for hypertension classification. “Higher” MAP was the primary exposure of interest in the case-control study, for which a threshold was not defined.

**Table 2.** Effect estimates for cohort studies investigating the association of hypertension with hepatocellular carcinoma risk.

| Country, Author (Year) | Participants, n | HCC Cases, n | Age at baseline, years | Sex (% male) | Relative risk measure<sup>†</sup> | Risk Ratio (95% Confidence intervals) |
| --- | --- | --- | --- | --- | --- | --- |
| UK, Ferreira (2020) (31) | 3927 | 16-19 | 39.2 (mean, non-DM)<br>54.9 (mean, DM) | 50 | aHR | 1.19 (0.95, 1.48) |
| France, Brichler (2019) (96) | 317 | 27 | 53 (median) | 82 | uHR | 2.91 (1.33-6.35) |
| Korea, Cho (2019) (75) | 826 | 86 | 52 (mean, NAFLD)<br>54 (mean, non-NAFLD) | 61 | aHR‡ | 1.38 (0.85-2.24) |
| China, Tan (2019) (76) | 4454 | 89 | 45.4 (mean) | 68 | aHR | 0.96 (0.60–1.53) |
| Taiwan, Hsu (2018) (53) | 27,820 | 802 | 48.1 (median) | 74 | uHR | 2.13 (1.85-2.45) |
| Hong Kong, Chan (2017) (80) | 270 | 11 | 43.6 (mean) | 75.2 | uHR | 2.33 (0.68-7.98) |
| US, Chayanupatkul (2017) (81) | 8539 | 317 | N/A | N/A | aHR‡ | 3.15 (1.02–9.75) |
| Korea, Kim (2017) (82) | 1696 | 24 | 50 (median) | 56.8 | aHR | 1.70 (0.71–4.05) |
| China, Gao (2013) (35) § | 370 | 122 | 54.7 (mean) | 86.9 | aOR‡ | N/A (p>0.1) |
| Taiwan, Hsu (2014) (88) | 210 | 35 | 52.8 (median) | 73.3 | uHR | 1.63 (0.74-3.60) |
| Taiwan, Chen (2008) (93) | 3931 | 187 | 45.9 | 59.3 | aHR | 0.45 (0.16–1.21) |

aHR, adjusted Hazards Ratio; uHR, unadjusted Hazards Ratio.
† Adjusted risk ratios are minimally adjusted for age and sex.
‡ Adjusted for age but not sex.
§ Case-control study using hospital controls.

Among 10 studies reporting hazards of HCC associated with HT, only three identified significantly increased risks, with two unadjusted and one adjusted for age. Another five studies reported an effect in the same direction, but effect sizes were not statistically significant. Adjusted HRs > 1 ranged from 1.19 to 1.70 and < 1 from 0.04 to 0.96. Adjustment for confounders was poor, with only four HRs minimally adjusted for age and sex.

### Associations of other comorbidities with hepatocellular carcinoma risk

Seven studies investigated the association of dyslipidaemia with HCC risk in CHB patients (Table 3). All studies reported reduced risks of HCC in participants with dyslipiaemia as compared to those without, however only one HR was statistically significant. Clinical definitions of dyslipidaemia were often not reported, and only four studies minimally adjusted for age and sex. Six studies investigated the association of obesity with HCC risk. Clinical definitions of obesity varied greatly, and out of four studies reporting increased risks of HCC with obesity, only one HR was statistically significant.

**Table 3.** Effect estimates for associations of other comorbidities and metabolic factors with hepatocellular carcinoma risk.

| Comorbidity | Relative risk measure<sup>†</sup> | Risk Ratio (95% Confidence intervals) |
| --- | --- | --- |
| **Dyslipidaemia** |  |  |
| UK, Ferreira (2020) (31)‡ | aHR | 0.77 (0.63, 0.95) |
| Korea, Cho (2019) (75) | uHR | 0.60 (0.26-1.39) |
| China, Tan (2019) (76) ‡ § | aHR | 0.67 (0.39–1.16) |
| China, Tan (2019) (76) ‡ ¶ | aHR | 0.49 (0.12–2.00) |
| US, Chayanupatkul (2017) (81) ‡ | uHR | 0.86 (0.38–2.00) |
| Taiwan, Hsu (2014) (88) | uHR | 0.34 (0.05-2.50) |
| Taiwan, Wu (2014) (55) ‡ ¶ | aHR | 0.87 (0.61-1.23) |
| **Obesity (BMI ≥ 30 kg/m<sup>2</sup>)** |  |  |
| France, Brichler (2019) (96) | aHR | 2.67 (1.04-6.84) |
| US, Chayanupatkul (2017) (81) | aHR†† | 2.21 (0.75–6.56) |
| **Obesity (BMI > 30 kg/m<sup>2</sup>)** |  |  |
| China, Tan (2019) (76) | aHR | 1.59 (0.73–3.44) |
| **Obesity (BMI ≥ 27.5 kg/m<sup>2</sup>)** |  |  |
| Netherlands, Brouwer (2015) (85) | uHR | 1.90 (0.60-6.20) |
| **Obesity (BMI ≥ 27 kg/m<sup>2</sup>)** |  |  |
| Taiwan, Chen (2013) (90) | uHR | 0.84 (0.44–1.60) |
| **Obesity (BMI ≥ 25 kg/m<sup>2</sup>)** |  |  |
| Korea, Lee (2016) (97) | uHR | 0.90 (0.17-4.62) |
| **Statin use** |  |  |
| UK, Ferreira (2020) (31) | aHR | 0.36 (0.19, 0.68) |
| Hong Kong, Yip (2020) (98) | aHR | 0.81 (0.73–0.90) |
| Hong Kong, Yip (2018) (78) | aHR | 0.52 (0.26–1.01) |
| **CVD** |  |  |
| Korea, Cho (2019) (75) – CVD | uHR | 1.46 (0.64-3.34) |
| US, Chayanupatkul (2017) (81) – IHD | uHR | 1.19 (0.39–3.65) |
| France, Mallet (2017) (83) - CVD | aHR | 0.57 (0.53–0.62) |
| Taiwan, Wu (2014) (55) - ACS | aHR | 0.66 (0.55-0.79) |
| Taiwan, Wu (2014) (55) - Cerebrovascular disease | aHR | 0.50 (0.40-0.64) |
| **Other** |  |  |
| Hong Kong, Yip (2020) (98) - Thiazolidinedione use | aHR | 0.57 (0.28-1.14) |
| Hong Kong, Yip (2020) (98) - Metformin use | aHR | 0.93 (0.85-1.02) |
| Hong Kong, Yip (2020) (98) - Sulphonylurea use | aHR | 1.19 (1.09-1.30) |
| Hong Kong, Yip (2020) (98) - Insulin use | aHR | 1.32 (1.19-1.46) |
| Hong Kong, Yip (2020) (98) - Aspirin/clopidogrel use | aHR | 0.76 (0.68-0.85) |
| Korea, Cho (2019) (75) - NAFLD | aHR | 1.67 (1.05-2.63) |
| France, Mallet (2017) (83) - Renal disease (off dialysis, no transplant) | aHR | 0.34 (0.28-0.41) |
| France, Mallet (2017) (83) - Renal disease (on dialysis, no transplant) | aHR | 0.36 (0.22-0.60) |
| France, Mallet (2017) (83) - Respiratory disease | aHR | 0.37 (0.33-0.42) |
| Taiwan, Yu (2017) (99) - Metabolic risk factors††† | aHR | HRs for obese or diabetic, obese and diabetic, and ≥3 metabolic risk factors were 1.29 (95% CI 0.92-1.81), 1.18 (0.16-8.54) and 2.61 (1.34-5.08), respectively, compared to the non-obese and non-diabetic reference group. |
| Taiwan, Wu (2014) (55) - COPD | aHR | 0.71 (0.59-0.87) |
| Taiwan, Wu (2014) (55) - Renal failure | aHR | 0.84 (0.68-1.04) |
| Taiwan, Chen (2013) (90) - Metabolic syndrome | uHR | 1.19 (0.63-2.26) |

aHR, adjusted Hazards Ratio; uHR, unadjusted Hazards Ratio; BMI, body mass index; CVD, cardiovascular disease; IHD, ischaemic heart disease; ACS, acute coronary syndrome; NAFLD, non-alcoholic fatty liver disease; COPD, chronic obstructive pulmonary disease.
† Adjusted risk ratios are minimally adjusted for age and sex.
‡ Defined specifically as hyperlipidaemia.
§ Defined specifically as hypertriglyceridaemia.
¶ Defined specifically as hypercholesterolaemia.
†† Adjusted for age but not sex.
††† Metabolic risk factors (obesity, diabetes, hypertriglyceridemia and HT), with exposure groups split into groups of 0, 1, 2 and ≥3 risk factors.

Three studies investigated the association of statin use with HCC risk in CHB. All studies reported HRs < 1, and two of these HRs were statistically significant. HRs reported in 5 studies for HCC risk associated with CVD varied, likely due to the variable definitions of CVD used across studies. Associations for other variables, including respiratory disease and renal disease, were reported by < 2 studies each.

## Discussion

Our meta-analysis suggests that DM is a risk factor for HCC in CHB infected individuals, with hazards of HCC > 20% higher in the presence of DM; however, we report significant between-study heterogeneity. This association did not materially change after restriction to studies adjusting for relevant confounders, but did suggest a favourable impact of DM treatment with metformin. Pooled effect estimates remained significant in sensitivity analyses. Few studies investigated other comorbidities, and some comorbidity search terms included in our systematic literature search returned few or no results. This highlights the need for future investigation of these comorbidities, as antiviral treatment cannot eliminate the risk of HCC entirely and therefore novel risk factors must be identified in order to inform interventions. Although EASL (7) and APASL (32) guidelines recognise this association, it is not currently consistently described in other recommendations (e.g. AASLD guidelines(33) do not list DM as a risk factor for HCC).

Some studies investigating comorbidities and their metabolic risk factors reported significantly reduced hazards of participants with these conditions as compared to those without. This association may be confounded by the requirement for treatment in secondary care, whereby CHB-infected individuals may be more likely to receive screening and antiviral treatment.

Findings from case-control and cohort studies were not consistent; whilst the majority of cohort studies reported increased risks of HCC associated with DM, case-control findings were inconsistent, and indeed three studies reported a significant reduction of HCC risks in association with DM. Explanations for such findings including confounding, selection bias associated with the study of hospital control groups that enrich for DM (34,35), and chance, especially in small studies (34–37).

Our findings are consistent with a previous meta-analysis (38); we provide a comprehensive review of all cohort studies and include a larger number of studies. We restricted to studies reporting HRs minimally adjusted for age and sex. However, adjustment for covariates and inclusion criteria varied considerably between studies, and this may explain some of the between-study heterogeneity. Substantial heterogeneity remained in sensitivity analyses restricted to studies adjusting for additional key confounders, as adjustment for confounders was variable within these studies and populations may not have been comparable. Although baseline age and sex characteristics were comparable across studies, there was variability regarding exclusion of those with additional comorbidities and those on antiviral treatment.

We noted variable definitions of DM, with some studies restricting investigation to type 2 DM whereas others included participants with unspecified DM. Risk factors for types 1 and 2 diabetes mellitus vary, and heterogeneity in DM definitions could therefore contribute to variable study populations and outcomes. Global prevalence and incidence estimates for specific DM types do not exist, as distinguishing between types often requires expensive laboratory resources that are not available in many settings. However most cases of type 1 diabetes are found in Europe and North America, and the large majority of studies included in this systematic review and meta-analysis were conducted in Asian countries (39).

It is possible that varied lengths of follow-up also contributed to between-study heterogeneity, although HRs did not significantly vary with length of follow-up in sensitivity analysis. This is because cancer is a chronic disease with a slow development, and preclinical disease can be present for many years before clinical manifestation; follow-up times < 10 years may be insufficient to detect HCC outcomes. We were unable to provide effect estimates across most potential patient subgroups because the subgroups contained small numbers of studies, putting subgroup analyses at greater risk of chance findings as well as being subject to the influence of multiple testing.

The association we report in this meta-analysis is weaker than those observed in patients with chronic HCV infection. In previous studies of individuals with chronic HCV infection, risk of HCC was elevated ∼2-fold in the presence of DM (40,41). Previous studies also report increased risks of DM in HCV-infected individuals as compared to non-infected individuals (76–79). However, this is likely due to the various extra-hepatic manifestations of HCV which are not present in HBV infection.

In sensitivity analysis restricted to studies adjusting for cirrhosis, the observed association of DM with HCC was attenuated towards the null. This may be explained by a confounding of the association by cirrhosis, accounted for by an independent association of cirrhosis with both DM and HCC, and the absence of cirrhosis from the causal pathway that associates DM with HCC. However, if cirrhosis is located along this causal pathway, then it can be characterised as a mediator rather than a confounder. If cirrhosis is a mediator then adjusting for it would be incorrect.

Past studies support a positive association of DM with HCC risk in non-CHB patients (42–44), and aetiological investigation has suggested that DM can lead to cirrhosis and thereby HCC via fatty liver disease (45), as a result of accumulation of fatty acids causing oxidative stress driving inflammation and tissue necrosis (46,47), and longer-term fibrosis and cirrhosis, thereby increasing HCC risk. However alternative pathways causally associating DM with HCC have been suggested, including increased hepatocyte proliferation induced by hyperinsulinaemia (48,49) and production of pro-inflammatory cytokines that increase cell survival via apoptosis inhibition (50–52). It is possible that multiple disease pathways associating DM with HCC operate simultaneously. Elucidation of the aetiological mechanisms underpinning this association will inform future epidemiological studies and disease management. Characterising the impact of glycaemic control on HCC risk is also an important question for future research.

Three studies adjusted for metformin use (53–55), and in sensitivity analysis restricted to these studies, the association between DM and HCC remained significant but was attenuated towards the null. It is not known the extent to which this is a result of glucoregulation by metformin, accomplished by inhibition of hepatic gluconeogenesis and improvement of insulin sensitivity in tissues leading to reduced oxidative stress in the liver (56), and/or a direct impact of metformin in reducing cancer risk via regulation of cellular signalling. Evidence from observational studies (57–59) and randomised controlled trials (RCTs) (60) supports a protective effect of metformin against the development and progression of cancer in diabetic individuals. There is also some RCT evidence for protective effects of metformin against progression of certain cancer types in non-diabetic individuals (61) although this is not consistent. Multiple large-scale phase III RCTs are currently underway (62–65) and will provide further information regarding the roles of DM and metformin in cancer development.

We included all studies investigating the association of comorbidities with risk of CHB progression to HCC that minimally adjusted for age and sex in order to provide a comprehensive review of available evidence. However, few studies investigated non-DM comorbidities, preventing meta-analysis for these comorbidities. Additionally we were unable to restrict our meta-analysis of DM and HCC to studies adjusting for confounders other than age and sex, as few studies minimally adjusted for all relevant factors. Publication bias may influence the outcome, as we restricted our search to peer-reviewed literature, and studies that do not report an association of DM with HCC may be less likely to be published. Our results may not be generalisable to the global CHB population, as there were a limited number of studies from non-Asian countries. The lack of studies from any African countries is of concern, given that the region carries both the highest HBV prevalence (3) and largest mortality burdens for cirrhosis and HCC (66,67).

Our finding that DM is a risk factor for HCC in CHB-infected individuals suggests that enhanced cancer surveillance may be justified in patients with CHB and DM to enable early detection and treatment. Improvements in guidelines could help to inform more consistent approaches to risk reduction. After adjustment for metformin use, this association remained significant, but was attenuated suggesting a potential benefit of metformin that warrants further study. Ongoing investigation is required in order to identify and characterise risk factors for HCC, to extend these analyses to diverse global populations, and to elucidate disease mechanisms in order to inform prevention, screening and therapeutic intervention.

## Data Availability

N/A, meta-analysis

## Acknowledgements

CC acknowledges doctoral funding from GlaxoSmithKline.

